# Path to Prevention Platform Trial in Early-Stage Neuronal Alpha-Synuclein Disease: Study Design

**DOI:** 10.64898/2026.11.04.26367309

**Authors:** Ruth B. Schneider, Christopher S. Coffey, Catherine M. Kopil, Maggie M. Kuhl, Cora Allen-Savietta, Michael C. Brumm, Seung Ho Choi, Amy Crawford, Dixie Ecklund, Kimberly Fabrizio, Tatiana Foroud, Frederic S. Goldstein, Cornelia Kamp, Karl Kieburtz, David-Erick Lafontant, Danielle N. Larson, Nijee Luthra, Nicola Pavese, Andrew Siderowf, Craig E. Stanley, Caroline M. Tanner, Thomas F. Tropea, Barbara Wendelberger, Tawny Willson, Sohini Chowdhury, Kenneth Marek, Tanya Simuni, the Path to Prevention Investigators and Parkinson’s Precision Medicine Initiative

**Affiliations:** Department of Neurology, University of Rochester, Rochester, NY 14642, United States; Department of Biostatistics, University of Iowa, Iowa City, IA 52246, United States; The Michael J. Fox Foundation for Parkinson’s Research, New York, NY 10163, United States; Berry Consultants, LLC, Austin, Texas 78746, United States; Institute for Neurodegenerative Disorders, New Haven, CT, 06510, United States; Department of Medical and Molecular Genetics, Indiana University School of Medicine, Indianapolis, IN, 46202, United States; The Parkinson’s Precision Medicine Initiative Community Advisory Board, New York, NY, 10163, United States; Department of Neurology, Northwestern University, Chicago, IL, 60611, United States; Department of Neurology, University of California San Francisco, San Francisco, CA, 94158, United States; Clinical Ageing Research Unit, Newcastle University, Newcastle upon Tyne, NE1 7RU, United Kingdom; Department of Neurology, University of Pennsylvania, Philadelphia, PA, 19104, United States

**Keywords:** [6] Parkinson’s disease, disease prevention, platform trial, neuronal synuclein disease

## Abstract

The development of biologically targeted therapeutics (BTTs) that slow Parkinson’s disease progression has been hampered by a reliance on testing interventions in clinically defined populations with already advanced pathobiology. Publications on the biological definition and classification of neuronal ⍰-synuclein disease (NSD), and the development of the NSD integrated staging system (NSD-ISS), have generated a framework for identifying individuals with NSD prior to the emergence of a clinically defined syndrome.

Path to Prevention (P2P) is a phase 2A, randomized, double-blind, placebo-controlled platform trial designed to assess the safety and efficacy of interventions in an early-stage NSD population. The trial is embedded within the observational Parkinson’s Precision Medicine Initiative (PPMI), a structure that enables recruitment of participants from the PPMI cohort, leverages existing operational infrastructure, and utilizes data from non-P2P enrolled PPMI participants in the Bayesian statistical modeling. P2P will enroll participants at least 60 years of age with NSD Stage 2B (abnormal ⍰-synuclein, dopaminergic dysfunction, and prodromal signs or symptoms without functional impairment) without a clinically defined diagnosis of PD or a related disorder. As a platform trial, P2P will enable the testing of multiple interventions under a Master Protocol at approximately 35 sites across North America and Europe. Interventions are selected from industry-submitted applications based on biological relevance of the targets, and on the adequacy of the safety data appropriate to the target population. Each treatment regimen will have 125 participants on active treatment and at least 125 shared placebo control participants. Participants will remain on treatment for a maximum of 144 weeks or until 96 weeks since the randomization of the last participant. P2P has two primary endpoints, 1) rate of change in mean putamen specific binding ratio from baseline through follow-up and, 2) rate of change in the MDS-UPDRS Part III motor examination from baseline through follow-up or initiation of dopaminergic treatment. A range of secondary and exploratory biomarker-focused endpoints will be assessed. The protocol was developed with input from multiple stakeholders, including people with early-stage NSD, biopharmaceutical industry leaders, and regulatory authorities. P2P is the first trial to test biologically targeted assets in biologically defined prodromal participants utilizing an efficient platform design, which will provide the field with tools to expand therapeutic development, and enable intervention at earlier stages of neurodegeneration. P2P is scheduled to begin enrollment in December 2026 and is committed to data sharing and broad dissemination of learnings from the study.

## Introduction

The single largest unmet need in Parkinson’s disease and related disorders is for biologically targeted therapeutics (BTTs) that slow and ultimately prevent disease progression. In the absence of a biological definition for Parkinson’s disease, potential BTTs have historically been tested in clinically defined populations. However, by the time a clinical diagnosis of Parkinson’s disease can be established, the underlying pathobiology is already fairly advanced,^1–3^ and may be too advanced for successful disease modification. Intervening before the onset of a clinically defined syndrome may be a more successful strategy for slowing disease progression and preventing disability. While Parkinson’s disease has characteristic prodromal features (including hyposmia, REM sleep behavior disorder, dysautonomia, and mood impairment) that enable the identification of individuals before a clinical diagnosis, the lack of understanding of the natural history of the earliest (pre-diagnosis) stages of disease and the absence of a clear regulatory-acceptable definition for prodromal Parkinson’s disease has prohibited the conduct of clinical trials of BTTs in this population.

However, with recent advancements, we are now prepared to initiate such trials. The development of reliable biomarkers for α-synuclein pathology has led to the emergence of biological disease frameworks, including the neuronal α-synuclein disease integrated staging system (NSD-ISS) and SynNeurGe, that define disease based on the presence of α-synuclein pathology.^4,5^ These frameworks have enabled the identification of individuals much earlier in the disease course, prior to the emergence of the clinically defined syndromes of Parkinson’s disease and dementia with Lewy bodies, and empowered therapeutic development. The Michael J. Fox Foundation for Parkinson’s Research (MJFF)-sponsored Parkinson’s Precision Medicine Initiative (PPMI), which has been investigating clinical, imaging, biological, and digital progression in Parkinson’s disease since 2010,^6^ has helped shape understanding of prodromal Parkinson’s disease and pioneered scalable methods for identifying biologically at-risk individuals for research participation.^7^ Lastly, the landscape of BTTs currently in development is rich and includes, among others, α-synuclein-^8,9^, inflammation-^10,11^, and gene-targeted approaches.^12–15^

These developments have directly enabled the Path to Prevention (P2P) trial, which will test the safety and efficacy of multiple BTTs for the treatment of biologically defined prodromal Parkinson’s disease. P2P will be embedded within PPMI, thereby enabling the use of existing study infrastructure and recruitment of participants directly from PPMI. P2P will use an innovative platform design that will enable the testing of multiple BTTs under a master protocol with efficient use of shared placebo controls and unifying study infrastructure. As the first interventional clinical trial in the field to examine the potential for BTTs to slow progression in a biologically defined prodromal Parkinson’s disease population, P2P is also a learning study intended to inform the broader field and advance the development of BTTs in this population. Key anticipated learnings relate to the paradigm for participant selection and recruitment, measurement of disease progression in the context of an early interventional study, endpoint performance in a population pre-clinical diagnosis, and advancement of regulatory science. Here, we describe the design of P2P and discuss key trial design considerations. The companion manuscript by Wendelberger et al describes the modeling that informed the design of the trial and statistical considerations.

### Study Design

P2P is a multi-center, randomized, double-blind, placebo-controlled platform trial designed to evaluate the safety and efficacy of multiple investigational products for the treatment of early-stage NSD (**Figure 1**). While there are other platform trials in Parkinson’s disease,^16–18^ all are targeting clinically defined Parkinson’s disease, setting P2P apart as the only platform trial focused on early intervention and secondary prevention of clinically defined disease in a biologically-defined prodromal population. Similarly, while there are other examples of ongoing or soon-to-be initiated prevention trials testing investigational drugs (**Table 1**), all are targeting populations with REM sleep behavior disorder, and this will be the first study to enroll based on NSD status.^19–22^

**Figure 1.**
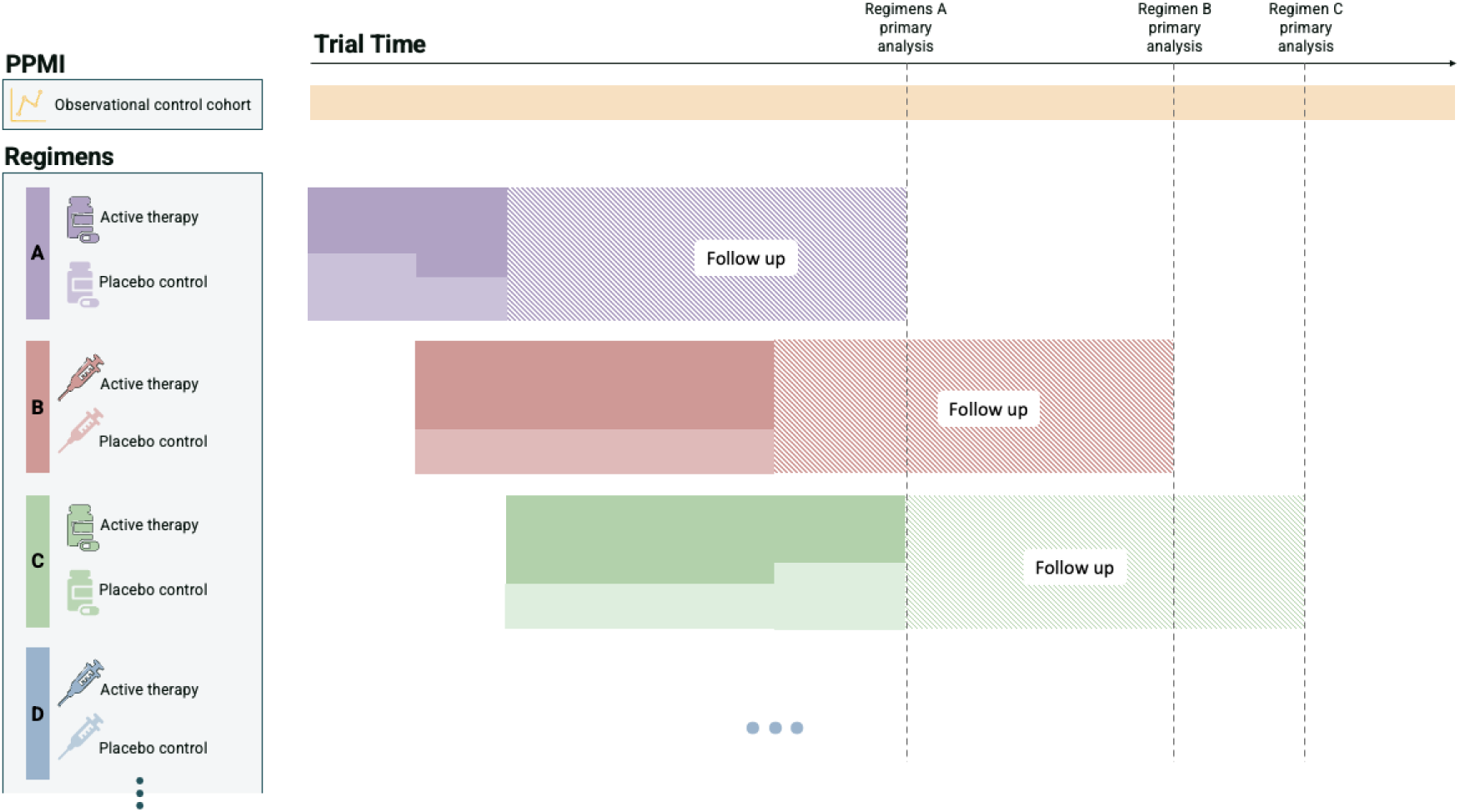
P2P platform trial design. This figure displays the design of P2P. Regimens may enter the platform trial at any time. Participants are randomized first to a regimen and then to active therapy or placebo, with the randomization ratio based on the number of actively enrolling regimens (as shown with shading).

**Table 1.** Landscape of PD Prevention Randomized Controlled Drug Trials.

|  | *P2P* | *PRISMS* | *SNT-4728* | *EITRS* |
|---|---|---|---|---|
| **Study Population** | NSD<sup>a</sup> Stage 2B | iRBD<sup>e</sup> | iRBD<sup>e</sup> | iRBD<sup>e</sup> |
| **Study Design** | Platform, double-blind | Two arms, double-blind | Two arms, double-blind | Two arms, double-blind |
| **Study Phase** | Phase 2 | Phase 2 | Phase 2 | Phase 2 |
| **Randomization** | k:1 | 1:1 | 3:1 | 1:1 |
| **Sample Size** | 125 per arm + shared placebo | 54 per arm | 40 total | 71 per arm planned; 27 total enrolled |
| **Treatment(s)** | Multiple; launching with c-Abl inhibitor risvodetinib | TNFα inhibitor adalimumab | Anti-inflammatory SNT-4728 | Antioxidant idebenone |
| **Treatment Duration** | Minimum 96 weeks | 96 weeks | 12 weeks | 5 years |
| **Primary Endpoint(s)** | Δ DAT SPECT mean putamen SBR<sup>b,c</sup> and Δ MDS-UPDRS part III score<sup>d</sup> | Δ FDG PET<sup>f</sup> Parkinson Disease Related Pattern<sup>c</sup> | Δ translocator protein PET-measured striato-cortical microglia activation | Phenoconversion to PD, DLB or MSA |
| **Sponsor** | Michael J. Fox Foundation | Yale University | Syntara | Ruijin Hospital |
| **Current Status** | Not Yet Recruiting | Recruiting | Active, not recruiting | Active, not recruiting |

This list may not represent the full landscape of PD prevention drug trials in a rapidly evolving field. <sup>a</sup>Neuronal synuclein disease (NSD); <sup>b</sup>Dopamine transporter single-photon emission computed tomography (DAT SPECT), specific binding ratio (SBR); <sup>c</sup>From baseline through follow-up; <sup>d</sup>From baseline through follow-up or the initiation of dopaminergic treatment; <sup>e</sup>idiopathic REM sleep behavior disorder (iRBD); <sup>f</sup>18-flurodeoxyglucose (FDG) positron emission tomography (PET)

P2P will take advantage of the robust existing PPMI infrastructure to test multiple investigational products and targets of interest in participants with early-stage NSD at approximately 35 sites across North America and Europe. A Master Protocol provides the overall framework for the trial and enables the testing of multiple investigational products both simultaneously and sequentially. A regimen consists of an investigational product and matched placebo control and each regimen is described in a Regimen Specific Sub-Protocol (RSSP) that dictates procedures, assessments, and testing unique to that investigational product. In a two-step randomization process, participants are randomized first in an equal manner to one of *K* actively enrolling regimens and then in a *K*:1 manner to active treatment or placebo control within the regimen. The first step (regimen assignment) is unblinded and the second step (treatment assignment) is blinded. Each regimen will have approximately 125 participants on active treatment and approximately 125 shared placebo control participants; the number of shared placebo controls will depend on the number of prior and actively enrolling regimens. Participants will remain on treatment for a maximum of 144 weeks or until 96 weeks since the randomization of the last participant. The Master Protocol is provided in the **Supplementary Materials**.

### Advantages of a Platform Trial

Platform trials like P2P have many operational efficiencies. With traditional clinical trial designs, the number of treatment arms and the trial duration are prespecified and usually limited. Moreover, trial infrastructure must be assembled prior to study initiation and then disassembled at the conclusion. In contrast, platform trials can test multiple investigational products over time (either simultaneously or sequentially) under the same master protocol using the same trial infrastructure and have no defined end date, enabling investigational products to be added as they become available. The use of the same sites, investigators, coordinators, and procedures provides both cost and time advantages that may support faster and more efficient decision-making regarding whether or not to move therapeutic research forward.^18^ Platform trials also enable uniform clinical, biological, and neuroimaging data acquisition across the testing of multiple investigational products, which minimizes the need to train staff on new procedures each time a new regimen is added. Beyond operational efficiencies, platform trials have certain statistical efficiencies, as they enable sharing of placebo controls across different treatment arms and enable learnings about endpoint behavior to be incorporated into future iterations of the Master Protocol, which is important given the novel study population.

### Placebo Considerations in a Platform Trial

Placebo effects are a well-recognized challenge in Parkinson’s disease clinical trials^23^ and require special consideration in platform trials, where multiple agents are tested against a shared placebo control group. P2P will consider testing agents with oral, intramuscular, and intravenous routes of administration, but not more invasive routes of administration (e.g., intrathecal, neurosurgical) given safety and feasibility considerations. In P2P, each regimen will include a placebo control that is matched to the investigational product associated with that RSSP based on formulation, route of administration, and dosing regimen.

P2P will have multiple types of controls. For analysis of a given treatment regimen, placebo data across regimens will be shared to produce a single placebo control dataset for analyses. The sharing of placebo between regimens has clear statistical advantages and enables modeling of differential responses to placebo (e.g., intravenous versus oral). As a perpetual platform trial, there may be substantial time gaps between enrollment in the first arm and enrollment in the last arm. The use of Bayesian modeling with hierarchical borrowing allows for heterogeneity across individual baseline values and individual slopes, as well as heterogeneity across regimens due to minor differences in the inclusion/exclusion criteria or mode of administration and will partially mitigate the impact of temporal and placebo differences. See Wendelberger et al for more on the use of dynamic borrowing.

### Leveraging PPMI Infrastructure for Operational Success

PPMI has a robust infrastructure, which is critical to the success of P2P. Importantly, PPMI will enable the enrollment of an engaged and biologically characterized population.^7^ Additionally, P2P will use existing PPMI Cores, including the Data Management, Statistics, Site Management, Participant Engagement, Recruitment, Imaging, and Biorepository Cores, to recruit participants, support retention, and collect and analyze trial data.^7^ Lastly, after participants complete their participation in P2P, they will be encouraged to continue follow-up within PPMI, providing valuable long-term follow-up data. Leveraging the PPMI infrastructure for P2P is an efficient use of resources, ensures the availability of essential expertise in the standardization of biomarker collection and processing, and supports harmonization with PPMI.

### Study Governance

The MJFF-sponsored P2P trial is governed by a steering committee comprised of leading Parkinson’s disease researchers with representation of individuals with lived experience. In addition to a lead Principal Investigator (PI; TS), each RSSP has a PI. The sponsor and its operational teams are responsible for trial conduct, obtaining regulatory approval, and contracting while the industry partner provides investigational product and matching placebo as well as investigational product-related documentation, and is closely involved in the development of the RSSP. Each RSSP has a working group that includes representatives from industry partners who work in close collaboration with the P2P team to ensure appropriate study design, provide operational input, and facilitate on-going exchange of information.

### Study Population

Eligible participants are those who are enrolled in PPMI, meet criteria for NSD Stage 2B, are at least 60 years of age, do not have a clinical diagnosis of Parkinson’s disease, dementia with Lewy bodies or another neurodegenerative disease, and are not on dopaminergic therapy or a cholinesterase inhibitor. NSD Stage 2B participants are identified based on the presence of 1) neuronal α-synuclein as determined in this study by a positive cerebrospinal fluid α-synuclein seed amplification assay (SAA) collected and analyzed under standardized conditions (Amprion, San Diego, CA, USA),^24^ 2) dopamine dysfunction as determined by dopamine transporter single-photon emission computed tomography (DAT SPECT) lowest age- and sex-adjusted putamen specific binding ratio (SBR) < 75%,^25^ and 3) clinical symptoms or signs without functional impairment as determined by motor, cognitive, and other non-motor clinical anchors (**Figure 2**).^26^

**Figure 2.**
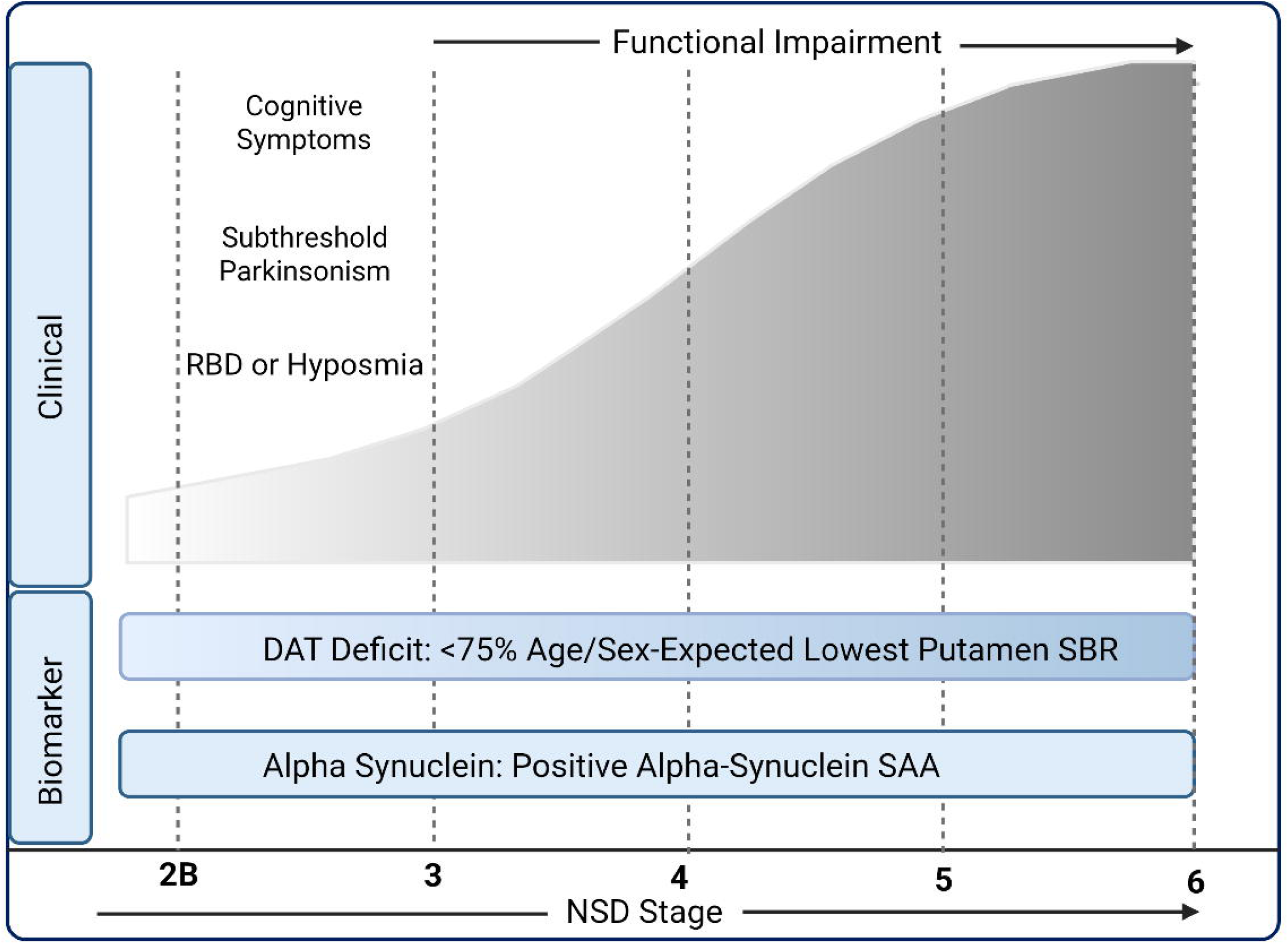
This figure shows the criteria for NSD Stage 2B. NSD Stage 2B is defined by the presence of a positive alpha-synuclein seed amplification assay, DaT deficit, and motor, cognitive, and other non-motor clinical anchors.

### Rationale for NSD-based Definition of the Study Population

Enrollment based on biological disease definition and staging has many advantages in prodromal cohorts. One, it ensures a homogeneous study population, defined by the presence of neuronal α-synuclein and dopamine dysfunction. In comparison, clinically defined prodromal cohorts are quite heterogeneous. While α-synuclein SAA assays have a high rate of positivity in RBD, up to 15% of individuals have a negative assay,^27–30^ and the positivity rate in older adults with hyposmia is approximately 50%.^31^ The necessity of biologically characterizing prodromal cohorts was further highlighted in a recent analysis of PPMI data, which demonstrated that 28% of RBD participants and 13% of hyposmic participants who were given diagnoses of clinically defined syndromes did not have the expected biological profile of synuclein pathology and dopamine dysfunction.^32^ Two, including a requirement for dopamine dysfunction in addition to α-synuclein enriches for individuals more likely to develop clinically defined syndromes. In the Parkinson Associated Risk Syndrome (PARS) study, 48% of older adults with median age of 65 years had a positive α-synuclein SAA assay, yet the majority did not have DAT deficit at baseline and did not develop DAT deficit over a median of 4 years of follow-up.^31^ However, the majority of participants with a positive α-synuclein SAA assay *and* DAT deficit at baseline developed parkinsonism during follow-up. Three, enrollment based on NSD Stage 2B status provides a clear and reproducible definition to replace the prior poorly defined term of prodromal disease. Lastly and essentially important, NSD Stage 2B demonstrates progression within a typical clinical trial timeframe; 21% of stage 2B PPMI participants without a clinical diagnosis progressed to functional impairment at year 1, 34% at year 2, and 43% at year 3.^33^

While earlier stage NSD populations (Stages 1-2A, individuals with a positive α-synuclein biomarker but no evidence of dopaminergic deficit and/or nor clinical symptoms or signs) may be reasonable for inclusion in future trials, at the present time, sufficient longitudinal data does not exist to inform understanding of progression and enrichment in these earlier stage populations.^34^ Lastly, later stages (Stages 3-6) are defined by the presence of functional impairment and the majority of participants in these stages are expected to have a clinically defined diagnosis.^4^ However, even among NSD Stage 2B there is heterogeneity of the clinical phenotype (see below). These phenotypic subgroups may display differential rates of progression and develop different clinical syndromes (e.g., Parkinson’s disease, dementia with Lewy bodies).^32^ Further work is needed to better characterize progression of these Stage 2B subgroups.

A minimum age of 60 years was established as the majority of PPMI participants are ≥ 60 years of age and sufficient data on NSD prevalence and progression are not yet available in a younger population. Consistent with the goals of P2P and the focus on early-stage disease prior to the onset of a clinically defined syndrome, individuals with a clinical diagnosis of Parkinson’s disease, dementia with Lewy bodies, or another neurodegenerative disease are excluded as are those who have received dopaminergic therapy or cholinesterase inhibitors within 90 days of the Baseline Visit. Individuals who have received drugs associated with drug-induced parkinsonism within 6 months of the Baseline Visit are excluded. Additional eligibility criteria, including safety-based exclusion criteria, are described in the **Supplementary Materials**.

### Recruitment and Enrollment of Early-Stage NSD Participants from PPMI

One of the major challenges of testing BTTs in prodromal Parkinson’s disease populations is the lack of an established paradigm for recruitment as these individuals do not typically have formal diagnoses (aside from RBD) or seek medical care. In recent years, the World Health Organization,^35^ Centers for Disease Control and Prevention,^36^ and American Academy of Neurology^37^ have all launched initiatives that promote brain health and the prevention of neurologic disease, and prevention-focused clinical models are emerging.^38^ Coupled with the increasing availability and accessibility of biomarkers, we anticipate that new recruitment paradigms will develop. However, at the present time, recruitment of biologically characterized prodromal populations into an interventional clinical trial would be impossible in the absence of a feeder cohort.

P2P participants will be recruited directly through PPMI, which has pioneered a staged recruitment paradigm for the identification of individuals with α-synuclein pathology.^39^ This staged recruitment paradigm begins with broad outreach to the community soliciting interest to remotely undergo smell testing and is followed by in-clinic screening of identified hyposmic individuals. Data have confirmed the utility of this approach for enriching for the presence of α-synuclein pathology and dopamine dysfunction.^40^ PPMI has remotely screened more than 133,000 individuals without PD, including over 25,000 with self-reported RBD and/or dream enactment behavior, since 2022. The clinical cohort includes more than 2800 classically defined prodromal participants with hyposmia, with RBD, and genetically at-risk for PD. As of July 2026, there are 2593 active prodromal participants in PPMI, of whom 1748 have NSD. Of these, 476 are NSD stage 2B; this includes participants with polysomnography-confirmed REM sleep behavior disorder (14%, n = 65), hyposmia (40%, n = 191), and hyposmia plus dream enactment behavior (43%, n = 207). Furthermore, 437 are at least 60 years of age, do not have a clinical diagnosis of Parkinson’s disease, dementia with Lewy bodies, or another neurodegenerative disease, and are not taking a dopaminergic medication. Outreach and screening will continue to meet P2P enrollment needs.

While we cannot accurately predict the proportion of these potentially eligible participants who will enroll in P2P, PPMI has set the foundation providing encouraging indicators. PPMI has pioneered the Return of Research Information (RORI) initiative, which enables interested PPMI participants to access their individual research results, including α-synuclein SAA status, DAT SPECT results, Movement Disorder Society-Sponsored Revision of the Unified Parkinson’s Disease Rating Scale (MDS-UPDRS) Part III scores, and smell test results.^41^ As of July 2026, 1983 PPMI prodromal participants have enrolled in RORI and 1771 have viewed their α-synuclein SAA results, indicating that interest in knowing one’s biological status is high. Additionally, qualitative interviews were conducted with prodromal PPMI participants regarding interventional study participation and these demonstrated strong interest.^7^ Complementing this work, a broad scale survey designed to assess perspectives on early intervention trials was administered to PPMI prodromal participants and will be published separately. P2P is thus well-positioned to successfully screen and enroll participants based on NSD status and to test biologically targeted interventions in this population.

### Study Endpoints

P2P is the first interventional trial to enroll participants based on NSD status. As a result, no regulatory precedent exists to inform the selection of endpoints. Outcome measures developed to assess clinical benefit in Parkinson’s disease may or may not be appropriate for use in an at-risk population. The U.S. Food and Drug Administration (FDA) defines meaningful clinical benefit as the effect an intervention has on how someone feels, functions, or survives.^42^ However, individuals with early-stage NSD may have minimal symptoms, and by definition, do not have functional impairment. This creates difficulties in the selection of an appropriate clinical endpoint. Thus, one of the goals of P2P is to generate the evidence needed to establish endpoints for this population.

### Primary Endpoints

We have done extensive modeling based on PPMI longitudinal data and arrived at the concept of using multiple primary endpoints which means that if a regimen is positive on one, it is considered a positive result. P2P will test whether treatment slows the rate of progression in a biomarker endpoint, DAT SPECT mean putamen SBR from baseline through follow-up, and a clinical endpoint, MDS-UPDRS Part III^43^ score from baseline through follow-up or initiation of dopaminergic treatment. The use of multiple primary endpoints allows for the simultaneous evaluation of biomarker and clinical progression, avoids the forced selection of a single primary endpoint when it is unknown which aspect of NSD will be more likely to respond to intervention, and provides more opportunities to demonstrate success, with a favorable outcome on either endpoint sufficient to establish efficacy.^44^ See Wendelberger et al for detailed rationale for endpoint selection and the underlying statistical modeling. In the eventual analysis of the data, we might identify other endpoints that show more sensitivity to change, and this is one of the learning aspects of the study. Endpoints assessing cognitive and neuropsychiatric progression are of essential importance and are included as exploratory endpoints, however, more data are necessary to inform sensitivity to change. We anticipate that P2P will inform the design and selection of future endpoints, both within the platform and in future trials in this population.

### Safety and Secondary Endpoints

Safety (as measured by treatment emergent adverse events (AEs) and serious adverse events (SAEs) for the active treatment versus placebo control within each RSSP), and secondary endpoints for feasibility and tolerability will also be assessed, as detailed in **Table 2**.

**Table 2.** P2P Study Endpoints.

| **Endpoint Category** | **Endpoint/Outcome Measure** | **Assessment Focus** |
|---|---|---|
| *Primary* | Rate of progression in the DAT SPECT mean putamen SBR<sup>a,b</sup> | Disease Progression |
|  | Rate of progression in the MDS-UPDRS Part III<sup>c</sup> | Disease Progression |
| *Safety* | All TEAEs<sup>d</sup> and SAEs<sup>e</sup> for the RSSP<sup>f</sup> active treatment arm vs. placebo control | Safety |
| *Secondary* | Ability to recruit and retain participants, and complete RSSP<sup>f</sup> study activities | Feasibility |
|  | Ability to complete an RSSP<sup>f</sup> on the assigned dose and treatment arm | Tolerability |
| *Key Exploratory* | Number who progress to NSD Stage 3 or higher | Clinical Progression |
|  | Rate of progression in the MDS-UPDRS total score<sup>c</sup> and Part I and II subscores<sup>a</sup> | Disease Severity |
|  | Number who develop new syndromes of MCI or dementia | Cognitive Outcomes |
|  | Rate of progression in the PDAQ-27<sup>a,g</sup> | Cognitive Outcomes |
|  | Time to progression milestones as defined by Brumm et al | Disease Progression |

<sup>a</sup>From baseline through follow-up; <sup>b</sup>Specific Binding Ratio (SBR);  <sup>c</sup>From baseline through follow-up or the initiation of dopaminergic treatment; <sup>d</sup>Treatment-emergent adverse events (TEAEs); <sup>e</sup>Serious adverse events (SAEs); <sup>f</sup>Regimen Specific Sub-Study Protocol (RSSP); <sup>f</sup>Penn Parkinson’s Daily Activities Questionnaire-27 (PDAQ-27)

### Key Exploratory Endpoints

A number of key exploratory endpoints will be assessed (**Table 2**). One key exploratory endpoint is the number who progress to NSD Stage 3 or higher. As noted above, approximately 20% of analyzed stage 2B PPMI participants progressed to functional impairment by year 1,^34^ suggesting that this will be an informative endpoint. However, progression to Stage 3 (and subsequent stages) is based on meeting clinical anchors (see **Supplementary Materials**), and valid concerns have been raised about the use of clinical anchors that are sensitive to dopaminergic medication.^45^ In a longitudinal analysis of PPMI data, 41% of participants who were Stage 4 at the last visit before starting dopaminergic medication reverted to an earlier stage at the first visit afterward.^34^ Notably, the same effect was not seen in Stage 2B participants and the majority progressed in staging following dopaminergic medication initiation, suggesting that initiation of dopaminergic medication is not a significant confounder in early-stage NSD. Concerns have also been raised regarding the lack of adequate representation of non-motor symptoms amongst the anchors and the insensitivity of the cognitive clinical anchors.^46^ Underscoring this, in a longitudinal analysis of PPMI data, 91% of those who progressed to Stage 3 did so only on motor anchors.^34^ Additional work is underway exploring novel approaches for clinical staging, which could change the NSD staging paradigm in the future.

In P2P, other key exploratory endpoints will bolster assessment of cognitive and other non-motor change. These endpoints include the rate of progression in MDS-UPDRS total score from baseline through follow-up or the initiation of dopaminergic treatment, rate of progression in MDS-UPDRS Part I and Part II subscores from baseline through follow-up, change in cognition as defined by the number of participants who develop new syndromes of mild cognitive impairment (MCI) or dementia, change in functional status as measured by the change in the Penn Parkinson’s Daily Activities Questionnaire-27 (PDAQ-27) from baseline through follow-up, and the time to progression milestones.^47^

### Exploratory Endpoints

A number of additional exploratory endpoints will also be assessed. As an example, time to phenoconversion to clinically manifest Parkinson’s disease or dementia with Lewy bodies will be included as an exploratory endpoint. While this endpoint has face validity as a clinical endpoint, it is inherently subjective. Moreover, annual phenoconversion rates are low; in an analysis of PPMI participants with RBD who were NSD Stage 2B or higher, only 33% phenoconverted over four years of follow up.^30^ Similarly, in an analysis of NSD Stage 2B PPMI prodromal participants, an estimated 4% phenoconverted at year one, 10% at year two, and 18% at year 3.^33^ Use of time to phenoconversion as a primary endpoint would therefore require prolonged trial follow-up.

An essential part of P2P is extensive biomarker characterization of the participants. Whole blood, urine, CSF, and skin biopsies will be collected for biomarker analysis using PPMI protocols, which will bolster biological understanding of the study population. For example, integration of biomarkers of neurodegeneration (e.g., amyloid, tau) will inform understanding of the influence of multi-pathology on treatment response. Similarly, biomarkers of different mechanistic pathways (e.g., neuroinflammation) will inform understanding of the influence of different etiological mechanisms on treatment response. The complete list of endpoints, including other exploratory endpoints, are included in the **Supplementary Materials**.

### Novel Endpoints for Biologically Targeted Therapeutics Trials in Early-Stage NSD

In trials for the earliest stages of Alzheimer’s disease, the FDA has demonstrated willingness, to consider changes on neuropsychological testing based on the size, breadth, and statistical robustness of the effect, as sufficient to establish meaningful clinical benefit when supported by biomarker evidence of effect.^48^ The European Medicines Agency is developing new guidelines to inform the development of treatments for Alzheimer’s disease that will consider appropriate endpoint selection in the early stages of Alzheimer’s disease and trial design in biologically defined pre-clinical Alzheimer’s disease.^43^ However, it is unclear how best to translate this to early-stage NSD, where individuals may meet criteria for Stage 2B based on motor symptoms, cognitive symptoms, or other non-motor symptoms (e.g., RBD, hyposmia) and exhibit different trajectories to different clinically defined syndromes. The MDS-UPDRS Part III is the best measure of motor performance currently available and has clinical meaningfulness.^49,50^ However, it has a floor effect^51^ and was not designed for use in a population without clinically defined Parkinson’s disease, raising questions regarding whether change in MDS-UPDRS Part III is an optimal primary endpoint.

Notably, P2P will provide additional evidence that may support the use of digital measures as clinical trial endpoints specifically suitable for this early NSD population and inform the development of novel outcome measures. Digital measures derived from digital health technologies may provide regulatory-acceptable^48^ sensitive measures of motor progression that can help to establish meaningful clinical benefit. P2P participants will wear a lumbar sensor (Axivity AX6) and a wrist sensor (Ametris ActiGraph LEAP) for 7 days every six months. This will enable passive, free-living data collection and capture of mobility and sleep measures using technically and analytically validated algorithms. Ongoing initiatives are collecting data to establish and validate digital measures for this context of use (disease progression monitoring) in early-stage NSD. Another opportunity lies in the development of novel fit-for-purpose patient-reported outcomes and clinician-reported outcomes, such as the Prodromal Synucleinopathy Rating Scale,^52^ which was modeled after the Clinical Dementia Rating (CDR) and has undergone preliminary cross-sectional validation in a large RBD cohort.^53^

### Regulatory Acceptance

There is no established regulatory pathway for studies testing therapeutics in prodromal synucleinopathies in part due to the lack of an operational definition for prodromal disease. Biologically defining prodromal disease based on NSD status fills this critical gap. In early stages of preparation for the study launch, the team engaged with the US and European regulatory authorities to initiate the dialogue on the acceptance of the definition of the population, platform study design, selection of the endpoints and other aspects of the study design. Both regulatory authorities provided valuable input in the context of Type C meetings in US and Trial Innovation Consultation with EMA, followed by country-specific Scientific Advise in the Netherlands and Germany. We received constructive feedback that was incorporated into the protocol. The regulatory authorities specifically highlighted the importance of biologically defining and staging the target population using the NSD-ISS framework in these early-stage development studies. The FDA Investigational New Drug (IND) application was filed using the NSD definition of the population and was granted “you may proceed status.” An EMA package will be submitted.

## Statistical Considerations

### Primary Analyses

The Bayesian primary analysis models estimate a difference in the rate of change of each primary endpoint from baseline through last visit (or, for the MDS-UPDRS Part III, through initiation of dopaminergic treatment). Each primary endpoint will be tested with equal priority with respect to active treatment superiority versus shared placebo controls. For both primary endpoints, a Bayesian repeated measures model of outcome will be used to compare the slope of active treatment versus a shared group of placebo and non-randomized controls. The model allows for heterogeneity across individual baseline values and individual slopes, as well as heterogeneity across regimens due to minor differences in the eligibility criteria or mode of administration. For each endpoint, if the Bayesian posterior probability of active treatment superiority (H_a_) is greater than or equal to a prespecified threshold of 0.90, the null hypothesis (H_0_) will be rejected, and the active treatment will be deemed superior to placebo control (i.e., less progression on active treatment) with respect to mean endpoint. This decision threshold of 0.90 has been calibrated via simulation to control the regimen-specific one-sided type I error at approximately 10% per endpoint per regimen (or 20% joint when considering both primary endpoints). It is hypothesized that an effective treatment will (1) decrease the progression rate and slow the decline in mean putamen SBR and/or, (2) decrease the progression rate and slow the increase in clinical symptoms as measured by the MDS-UPDRS Part III score.

The primary analyses for each analysis regimen will be based on the intent-to-treat (ITT) principle and include all participants randomized to the active treatment arm of the analysis regimen and all shared controls, regardless of the adherence to the assigned RSSP or duration of follow-up. Sensitivity analyses will be conducted using an ITT population that consists of participants randomized to the active treatment arm of the analysis regimen and (1) all shared P2P placebo controls (RSSP placebo, concurrent placebo, and non-concurrent placebo controls), (2) P2P concurrent controls (RSSP placebo, concurrent placebo), and (3) RSSP placebo controls. Lastly, we will also replicate the primary analyses using a per-protocol population. No interim efficacy or futility analyses are planned.

### Safety Analyses

Analyses to examine safety of each active treatment versus RSSP placebo controls will involve comparisons of the percentage of participants within each RSSP reporting treatment emergent AEs, treatment emergent SAEs, and unanticipated SAEs within each MedDRA (Medical Dictionary for Regulatory Activities) system organ class and preferred terms.

### Secondary Analyses

To examine tolerability, within each RSSP we will compare the percentage of participants on treatment vs regimen-specific placebo control group who complete the full RSSP on their initially assigned treatment regimen, regardless of reason for non-completion. Key feasibility metrics include enrollment timelines, retention, and visit compliance. Secondary analyses will include an ITT population consisting of all participants randomized to the active treatment arm or shared placebo control arm within an RSSP. Handling of key exploratory endpoints are as detailed in the **Supplementary Material.**

### Missing Data and Sensitivity Analyses

To assess the potential dependence of the results of the primary analysis on missing values, we will conduct a series of sensitivity analyses including comparison of RSSP treatment group to randomized placebo controls (excluding concurrent non-randomized controls), supplemented intention to treat analysis using PPMI data, and various multiple imputation methods.

### Sample Size Considerations

Each regimen will include approximately 125 participants on active treatment and approximately 125 shared placebo control participants as well as non-randomized controls from PPMI. To inform assumptions for simulating outcomes and the platform trial design, analyses were conducted using the PPMI Clinical prodromal cohort who met NSD Stage 2B criteria and additional eligibility criteria for the P2P Master Protocol (based on a download of the PPMI data from September 2025). See companion paper (Wendelberger et al) for additional details.

For a regimen, “power” is defined as the probability of demonstrating success on at least one of the primary endpoints. Regimens that enter the platform trial later are expected to see increased power relative to the first regimen due to the accumulation of shared, randomized placebo controls. In addition, the number of non-randomized controls from PPMI that will be available for each regimen analysis are unknown, and a range from 0 to 100 non-randomized control participants were considered in clinical trial simulation. Simulation results estimate power for the first three regimens between 73%-84% to detect treatment effects when both endpoints achieve 40% slowing of progression. Similarly, simulation results demonstrate 67%-77% power to declare success if there is truly a 40% or greater reduction in slope/progression of MDS-UPDRS Part III if there is at least a simultaneous 30% or greater reduction in mean putamen SBR. Finally, we have around 66%-79% probability of declaring success if there is truly a 40% or greater reduction in slope/progression on the DAT SPECT imaging endpoint and at least a 30% reduction on slope/progression on the MDS-UPDRS Part III endpoint. The simulations also demonstrated that the overall type I error probability (probability of meeting the criteria for success when there is no true reduction on either endpoint) was well controlled at 18%. Further details of Bayesian modeling for this study can be found in the companion paper.

### Selection of Investigational Products

Potential industry and academic partners are invited to submit letters of interest for testing an investigational product within P2P. As of July 2026, 28 letters of interest have been received. A recent review of phase 1-3 Parkinson’s disease drug trials registered on clinicaltrials.gov from 2015-2024, identified 165 trials of potential disease modifying agents^15,54^; the 28 agents submitted thus represents ∼17% of all BTTs currently in development. Thus far, interested industry partners have predominantly been those interested in early-stage NSD as an opportunity for label expansion for assets moving forward in Parkinson’s disease and those with assets with an initial indication for dementia with Lewy bodies or multiple system atrophy. A Therapeutic Evaluation Committee, comprised of international experts in pharmacology, biomarkers, and clinical trials design, is charged with the review and prioritization of potential investigational products for testing. The selection of interventions for P2P is based on biological relevance to an early-stage NSD population, safety data, appropriateness for testing in a platform trial infrastructure, and partnership readiness (e.g., availability of drug supply and willingness to comply with the P2P governance).^15^ As early-stage NSD is defined by the presence of neuronal α-synuclein pathology, α-synuclein provides a compelling target; however, other mechanistic pathways (e.g., inflammation) are also relevant at this stage of disease. Testing BTTs in a population without functional impairment that may never develop clinically defined disease, requires careful consideration of safety. Accordingly, only investigational products that have been tested in humans and have an established, acceptable safety profile will move forward. The Therapeutics Evaluation Committee also takes operational aspects, such as frequency and route of administration, into consideration. The final decision regarding the selection of investigational products for testing is made by the P2P Steering Committee. The first phase of P2P aims to test up to three compounds, the first of which is the oral, brain-penetrant c-Abl inhibitor risvodetinib.^55^ Other partnerships will be announced once the selection and contractual processes are completed.

### Schedule of Assessments

Following Master Protocol screening, RSSP screening, and the Baseline Visit, in-clinic visits will occur at Weeks 4, 12, 24, 48, 72, 96, 120, and 144 (and any other time points specifically required by a given RSSP). Remote visits will occur at Week 2 (telephone), Weeks 36, 60, 84, 108 and 132 (audio-video or telephone), and 30 days following RSSP treatment completion (telephone). Demographic variables include sex, age, race, ethnicity, and education status. Clinical assessments include the MDS-UPDRS, Hoehn & Yahr, modified Schwab & England, Epworth Sleepiness Scale, REM Sleep Behavior Disorder Screening Questionnaire, Scales for Outcomes in Parkinson’s Disease-Autonomic, Neuro-QOL, 15-item Geriatric Depression Scale, State-Trait Anxiety Inventory, Questionnaire for Impulsive-Compulsive Disorders in Parkinson’s Disease, UPSIT, Clinical Global Impression-Severity, Patient Global Impression-Severity, PDAQ27, MoCA, and a battery of neuropsychological tests. Physical assessments include height, weight, heart rate, and blood pressure (supine to standing). Safety assessments include clinical safety labs (whole blood and urine), electrocardiogram, and the Columbia-Suicide Severity Rating Scale. Imaging studies include DAT-SPECT and head MRI. Whole blood, urine, CSF, and skin biopsies will be collected in support of a broad array of biomarker analyses. Additional in-clinic visits may be completed as necessary for safety assessments, prior to initiation of dopaminergic therapy, if RSSP treatment is completed between visits, or in the event of early termination. The complete schedule of assessments can be found in the **Supplementary Materials**.

### Connectivity with PPMI and Innovation in Data Sharing

P2P, like PPMI, is committed to rapid, accessible data sharing and to the broad and timely distribution of study results. P2P participants will continue to contribute data to the PPMI data repository in real-time for the assessments and visits that align with the PPMI schedule of activities. To support data sharing, the schedule of assessments for the annual visits (Baseline, W48, W96, W144) has been constructed to align to the extent possible with the PPMI schedule of assessments. We have carefully considered potential or perceived risks of unblinding due to data sharing and concluded that these are exceedingly low. Notably, the shared data will not include any study identifiers. Moreover, any data specific to the P2P protocol or that carries risk of unblinding will be sequestered in accordance with industry standards until completion and publication of the relevant RSSP. **Table 3** summarizes novel aspects of the P2P study design and provides the rationale for key decisions.

**Table 3.** P2P Innovation.

| *Category* | *P2P Decision* | *Alternative* | *Rationale* |
|---|---|---|---|
| Study Population | Biologically defined, NSD Stage 2B | Clinically defined prodromal population | • Homogeneous population<br>• Enriches for those likely to progress within a clinical trial timeframe<br>• Reproducible, regulatory-accepted definition |
| Connectivity | Interventional study “embedded” in an observational study | Standalone trial | • Engaged and eligible population for recruitment<br>• Mature infrastructure<br>• Availability of data for modeling<br>• Non-randomized controls |
| Endpoints | Multiple endpoints | Single primary endpoint | • Simultaneous evaluation of biomarker and clinical progression<br>• More opportunities to demonstrate success |
| Study Design | Platform trial | Two-arm trial | • Simultaneous and sequential testing of agents as they become available<br>• Uniform data acquisition<br>• Statistical efficiencies |
| Risk Disclosure | Participants are informed of their biomarker status | No disclosure | • Consistent with the ethical principle of respect for persons |
| Data Sharing | Real-time contributions to PPMI data repository | Fully sequestered data | • Supports on-going scientific progress<br>• Low risk of unblinding due to data sharing |

### Study Timeline and Current Status

Study conceptualization began in 2022 with the launch of the formal, targeted planning phase of the study in 2024. The initial IND application was submitted to the FDA and opened October 2024. We present these timelines to highlight the substantial time and effort required to launch a platform trial in a novel population that has never previously been targeted for therapeutic interventions. The most time-consuming aspects of overall study preparation included conceptual design, stakeholders’ alignment, iterative statistical modeling adjusted based on an ever-expanding and maturing PPMI cohort dataset and selecting appropriate partnerships with pharma companies. Active study prelaunch activities started in December 2025. An updated Master Protocol and the first RSSP were submitted to FDA in April 2026. Approval from the WCG Institutional Review Board was received in August 2026. Submissions to the relevant regulatory authorities in the United Kingdom, Europe and Canada are anticipated later this year. The first U.S. participant is expected to be screened in the last quarter of 2026. We expect that learnings from the P2P planning phase will substantially facilitate and shorten timelines for the launch of future studies testing BTTs in early NSD.

## Conclusions

Therapeutic interventions that slow neurodegeneration are urgently needed and intervening earlier in the disease process may provide our best chance of success. P2P is a first-in-field platform trial that will take advantage of new biological frameworks and the success of PPMI to test the safety and early efficacy of the most promising BTTs in early-stage NSD. P2P includes multiple novel aspects. Learnings from the study will provide much needed tools for the broader research field and accelerate therapeutic development in populations at earlier stages of neurodegeneration.

## Supporting information

PPMI Study Group Authors

## Data Availability

Data used in the preparation of this article were obtained on 2026-07-27 from the PPMI database (https://www.ppmi-info.org/access-data-specimens/download-data), RRID:SCR_006431. For up-to-date information on the study, visit http://www.ppmi-info.org. Protocol information for The PPMI Clinical - Establishing a Deeply Phenotyped PD Cohort can be found on protocols.io or by following this link: https://dx.doi.org/10.17504/protocols.io.n92ldmw6ol5b/v2.

This analysis was conducted by the PPMI Statistics Core and used actual dates of activity for participants, a restricted data element not available to public users of PPMI data (Tier 4 Data).

Statistical analysis codes used to perform the analyses in this article are shared on Zenodo (10.5281/zenodo.23214203).

## Acknowledgements

PPMI – a public-private partnership – is funded by the Michael J. Fox Foundation for Parkinson’s Research and funding partners, including AbbVie, Alamar Biosciences, Aligning Science Across Parkinson’s (ASAP), Arrowhead Pharma, Arvinas, AskBio, BIAL, BioArctic, Biohaven, BlueRock Therapeutics, Bristol Myers Squibb, Calico Labs, Capsida Biotherapeutics, Critical Path Institute, DaCapo Brainscience, Denali, Edmond J. Safra Foundation, Eli Lilly, Gain Therapeutics, GE Healthcare, Genentech, GSK, Insitro, Johnson & Johnson Innovative Medicine, Lundbeck, Merck, Neumora, Neuron23, Novartis, Olink, Regeneron, Roche, Sanofi, Tenvie, UCB, Vanqua Bio, Voyager Therapeutics, The Weston Family Foundation.

## Supplementary material

Supplementary material is available at *Brain* online.

